# Using Coincidence Analysis to Identify Causal Chains of Factors Associated with Implementation and Optimization of Lynch Syndrome Tumor Screening Across Multiple Health care Systems

**DOI:** 10.1101/2024.02.22.24301533

**Authors:** Deborah Cragun, Zachary M Salvati, Jennifer L Schneider, Andrea N Burnett-Hartman, Mara M Epstein, Jessica Ezzell Hunter, Su-Ying Liang, Jan Lowery, Christine Y. Lu, Pamala A. Pawloski, Victoria Schleider, Ravi N Sharaf, Marc S Williams, Alanna Kulchak Rahm

## Abstract

**Purpose:** This study of multiple case study compared Lynch syndrome universal tumor screening (UTS) to understand multi-level factors that may impact the successful implementation of complex programs.

**Methods:** Data from 66 stakeholder interviews were used to conduct multi-value coincidence analysis (mv-CNA) and identify key factors that consistently make a difference in whether UTS programs were implemented and optimized at the system level.

**Results:** The selected CNA model revealed combinations of conditions that distinguish 4 optimized UTS programs, 10 non-optimized programs, and 4 systems with no program. Fully optimized UTS programs had both a maintenance champion and a positive inner setting. Two independent paths were unique to non-optimized programs: 1) positive attitudes and a mixed inner setting, or 2) limited planning & engaging among stakeholders. Negative views about UTS evidence or lack of knowledge about UTS led to a lack of planning and engaging, which subsequently prevented program implementation.

**Conclusion:** The model improved our understanding of program implementation in health care systems and informed the creation of a toolkit to guide UTS implementation, optimization, and changes. Our findings and toolkit may serve as a use case to increase the successful implementation of other complex precision health programs.

## INTRODUCTION

Lynch syndrome (LS) is the most common cause of hereditary colorectal and endometrial cancers.^1^ Effective reduction in morbidity and mortality from LS-associated cancers through increased cancer surveillance, prevention, and early treatment has been demonstrated.^2,3^ An estimated 1 in every 279 individuals in the US has LS; however, only about 2% are aware of it.^4,5^ Thus, an estimated 1 million individuals remain undiagnosed, highlighting a significant healthcare gap in initiation of cancer screening and prevention.^4^ One cost-effective and evidence-based approach to the identification of LS is universal tumor screening (UTS) for all patients newly diagnosed with colorectal and endometrial cancer.^6–8^ Briefly, a UTS program begins by testing tumors for mismatch repair deficiency (dMMR), a pathologic hallmark of LS. Individuals with dMMR (abnormal screen) are then referred for confirmatory germline genetic testing for a pathogenic germline variant in any one of four DNA mismatch repair genes (*MLH1*, *MLH2*, *MLH6*, and *PMS2*) or in rare cases, *EPCAM*. Implementation of UTS programs has been recommended since 2009 by the Evaluation of Genomic Applications in Practice and Prevention (EGAPP) Working Group^9^ and subsequently endorsed by multiple organizations.^10–17^ Furthermore, the identification of individuals with LS for cancer prevention is a Healthy People 2030 goal.^18^

There is no one “best” approach for implementing UTS programs, and successful UTS program implementation within health care systems has been challenging. It requires health system stakeholders to choose the best-fitting UTS approach and to coordinate across multiple service departments.^11,19^ Additional reported barriers to system-wide implementation include perceived high cost, lack of resources or personnel, and low levels of confidence by non-genetics clinicians in interpreting results.^10,11^ Given the complexity of health care systems, UTS implementation remains variable, if implemented at all.^20–22^

Few studies have systematically evaluated factors contributing to UTS program implementation or optimization (e.g., facilitators that improve consistency, efficiency, and high rates of genetics referrals and germline testing following abnormal screens).^20,23,24^ Purposeful use of implementation science frameworks and methods to understand variability and better guide UTS program implementation are warranted. The IMPULSS study^25^ employed the Consolidated Framework for Implementation Research (CFIR version 1.0) to characterize contextual factors that may serve as implementation barriers or facilitators over multiple levels of influence (i.e., individuals, inner setting, outer setting, intervention characteristics),^26,27^ and to understand contextual factors that may influence program optimization (i.e., greater efficiency and characteristics previously associated with higher rates of LS identification). Our systematic method of conducting cross-case comparisons of health care systems using process mapping and matrix heat mapping has been published previously^28^ describing how we categorized the outcome of each evaluated UTS program as optimized, non-optimized, or not-implemented, as well as the process for categorizing and selecting CFIR factors that may be relevant barriers or facilitators to implementation and optimization. Here we report the main findings of coincidence analysis (CNA) that allowed us to identify which barriers and facilitators distinguish between health care systems with no UTS, non-optimized UTS, and optimized UTS programs. CNA is an analytic method that employs a mathematical algorithm to build models and identify key factors that consistently make a difference in an outcome.^29,30^

## MATERIALS AND METHODS

We used data from process mapping and data matrix heat mapping^28^ to conduct CNA and identify key factors that consistently make a difference in the implementation and optimization of UTS programs across the multiple health care systems in the IMPULSS study (R01CA211723). Both Geisinger and Sutter Health IRBs approved this study, with Geisinger serving as the central IRB for all health care systems in the study except Sutter Health-Palo Alto Medical Foundation. The participating health care systems and overall study protocol are reported elsewhere.^25^

### Data collection, initial coding, and consolidation

Methods to collect and consolidate data have previously been described in detail.^28^ Trained qualitative researchers conducted semi-structured interviews with 66 genetic counselors, pathologists, oncologists, administrators, and others involved in 19 unique UTS programs/non-programs from 9 health care systems; creating the “cases” for this analysis. Interviews elicited information about UTS protocols/procedures (when relevant) and CFIR factors that may impact decisions to start a UTS program or how UTS programs were (or could be) implemented. Transcripts were coded, analyzed, and consolidated using process mapping to identify care gaps and/or inefficiencies within the UTS programs and data matrix heat mapping to categorize and visualize CFIR contextual factors. Process mapping revealed five components of UTS optimization (Table 1) which were quantified for each of the UTS programs to create an optimization score.

**Table 1.**
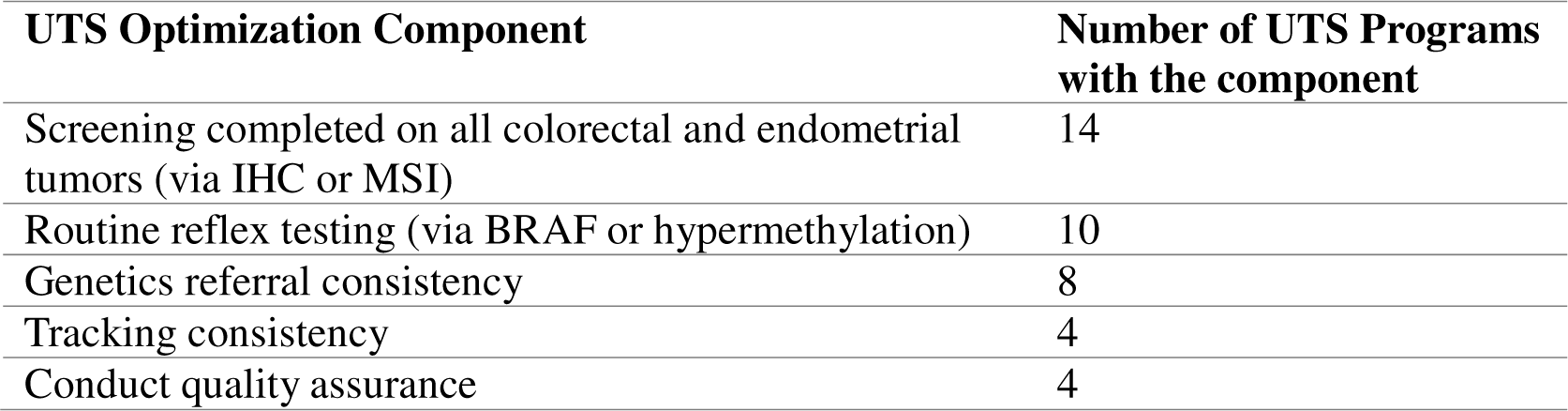
Program optimization by component and number of Programs.

| **UTS Optimization Component** | **Number of UTS Programs with the component** |
| --- | --- |
| Screening completed on all colorectal and endometrial tumors (via IHC or MSI) | 14 |
| Routine reflex testing (via BRAF or hypermethylation) | 10 |
| Genetics referral consistency | 8 |
| Tracking consistency | 4 |
| Conduct quality assurance | 4 |

UTS programs with all 5 optimization components present were categorized as fully optimized (n=4 cases) while those missing two to four components were categorized as non-optimized (n=11). One case was removed due to missing data, leaving 10 non-optimized programs for analysis. The remaining health care systems (n=4 cases) lacked a systematic UTS program, though some had other approaches to identify LS that were not systematic or universally implemented on tumors from all newly diagnosed colorectal and endometrial patients. A color-coded matrix heat map (supplemental figure 1) was created to summarize CFIR contextual factors hypothesized as possible difference makers for implementation and optimization based on prior literature and experience implementing UTS and other complex interventions in health care systems, including: 1) active use of external networks that support UTS (i.e., cosmopolitanism) or desire to keep up with other health care systems that previously adopted UTS (i.e., peer pressure); 2) evidence favoring UTS and its relative advantages; 3) concerns about UTS costs; 4) knowledge and attitudes about UTS; 5) an implementation champion; 6) inner setting characteristics (except for structural characteristics of the organizations); 7) evidence of a maintenance champion; and 8) stakeholder planning & engaging.

### Data Preparation for CNA: data calibration and reduction of data fragmentation

Prior to conducting CNA, the data matrix heat map was calibrated by assigning each case a numeric value that represented the prior color coding for each factor and outcome (Figure 1). For the implementation & optimization outcome, cases with no UTS program were assigned a value of 0, non-optimized programs were assigned a value of 1, and fully optimized programs were assigned a value of 2. After initial calibration, the number of factors and the number of factor values (referred to as either outcome values or conditions) were consolidated to reduce the high level of data fragmentation, which occurs when there are many possible conditional configurations and a relatively small number of cases (see supplemental material for details). Figure 1 shows the final consolidated and calibrated data with color coding for easier pattern recognition. Each row depicts a deidentified case (first column), showing the combination of CFIR factor values (i.e., conditional configuration) and respective implementation outcome values associated with that case. Unique cases from the same health care system are identified by the same number and differentiated by their letter.

**Figure 1.**
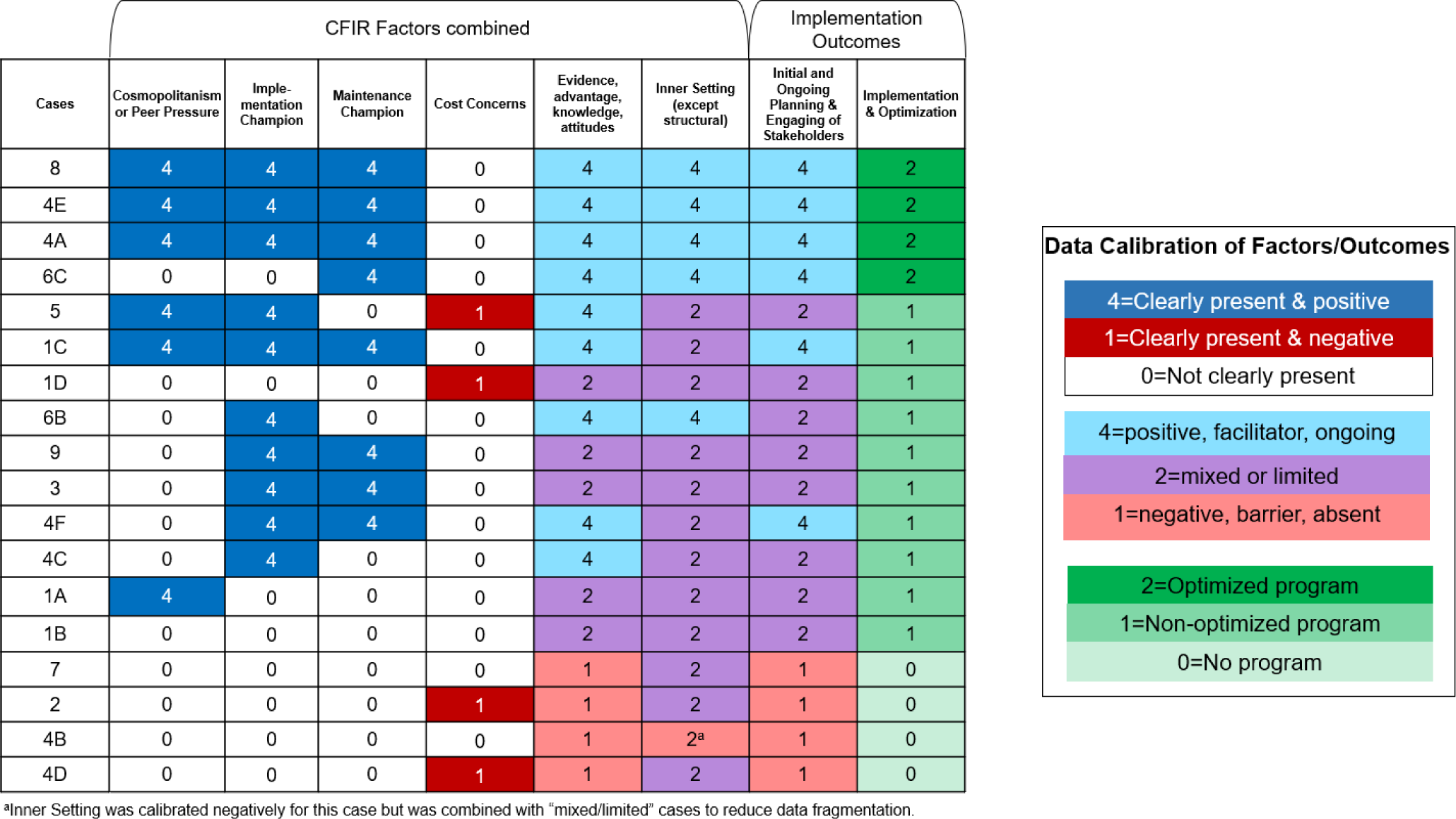
Final consolidated and calibrated data matrix of factors and implementation outcomes.

### CNA Modeling and Model Selection

The final calibrated data matrix and cna package in R statistical software were used to conduct multi-value coincidence analysis (mv-CNA). Factors were ordered according to Figure 2 and all hypothesized CFIR factors were permitted as potential difference makers for ‘Planning & Engaging’ or ‘Program Implementation & Optimization.’ This allowed for causal chains (if present) to be identified, whereby one or more values for ‘Planning & Engaging’ may be an intermediate outcome that makes a difference in whether a UTS program is implemented or optimized.

**Figure 2.**
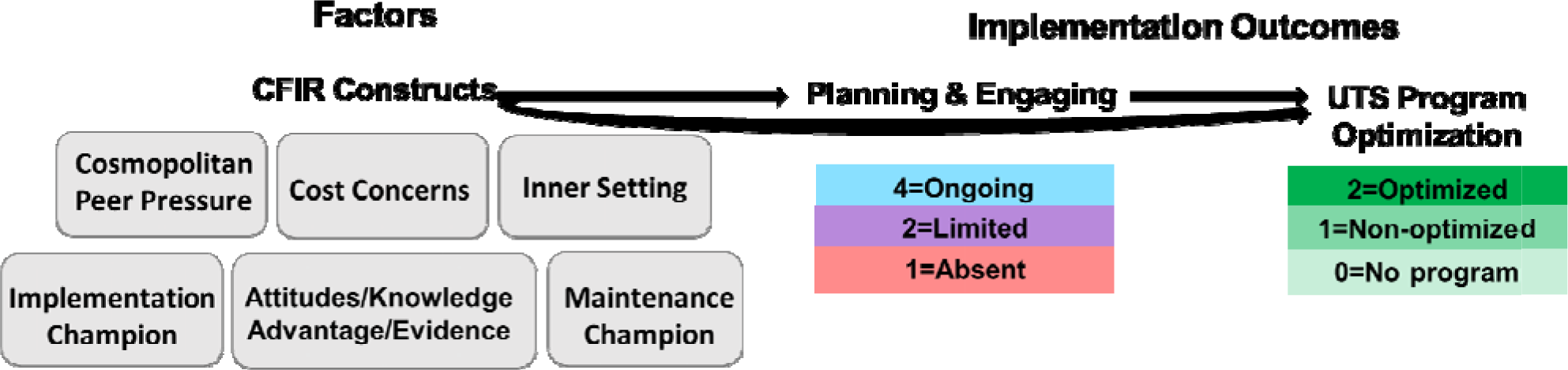
Hypothesized factors that may make a difference for implementation outcomes and the assigned values for each outcome.

The frscore package in R was used to analyze the data across various consistency and coverage thresholds between .75 and 1.0. The top-performing model solutions were reviewed and compared as described in the supplemental material. Briefly, members of the core analytic team (DC, ZS, AKR) reviewed coded interview data and pulled quotes relevant to factor/outcome relationships from the most robust models. Ultimately, all research team members reviewed the models and unanimously agreed upon the selected model presented here as the most applicable and useful based on the core team’s deep understanding of the underlying qualitative information and local systems knowledge held by team members.

## RESULTS

The model shown in figure 3 was selected from the top 9 most robust models generated and had a perfect consistency and coverage of 1.0. This selected model reflected the complexity inherent in UTS implementation and optimization, provided support for causal chains, included all outcome values, explained all cases, and was well supported by interview quotes. Conditions in this model were largely consistent with other robust but less complex models. The complexity detailed in this model was determined to be more helpful for achieving a key goal of the IMPULSS study, which was to develop a toolkit to address key factors that appear to contribute to successful planning & engagement among stakeholders as well as implementation & optimization of UTS programs.

**Figure 3.**
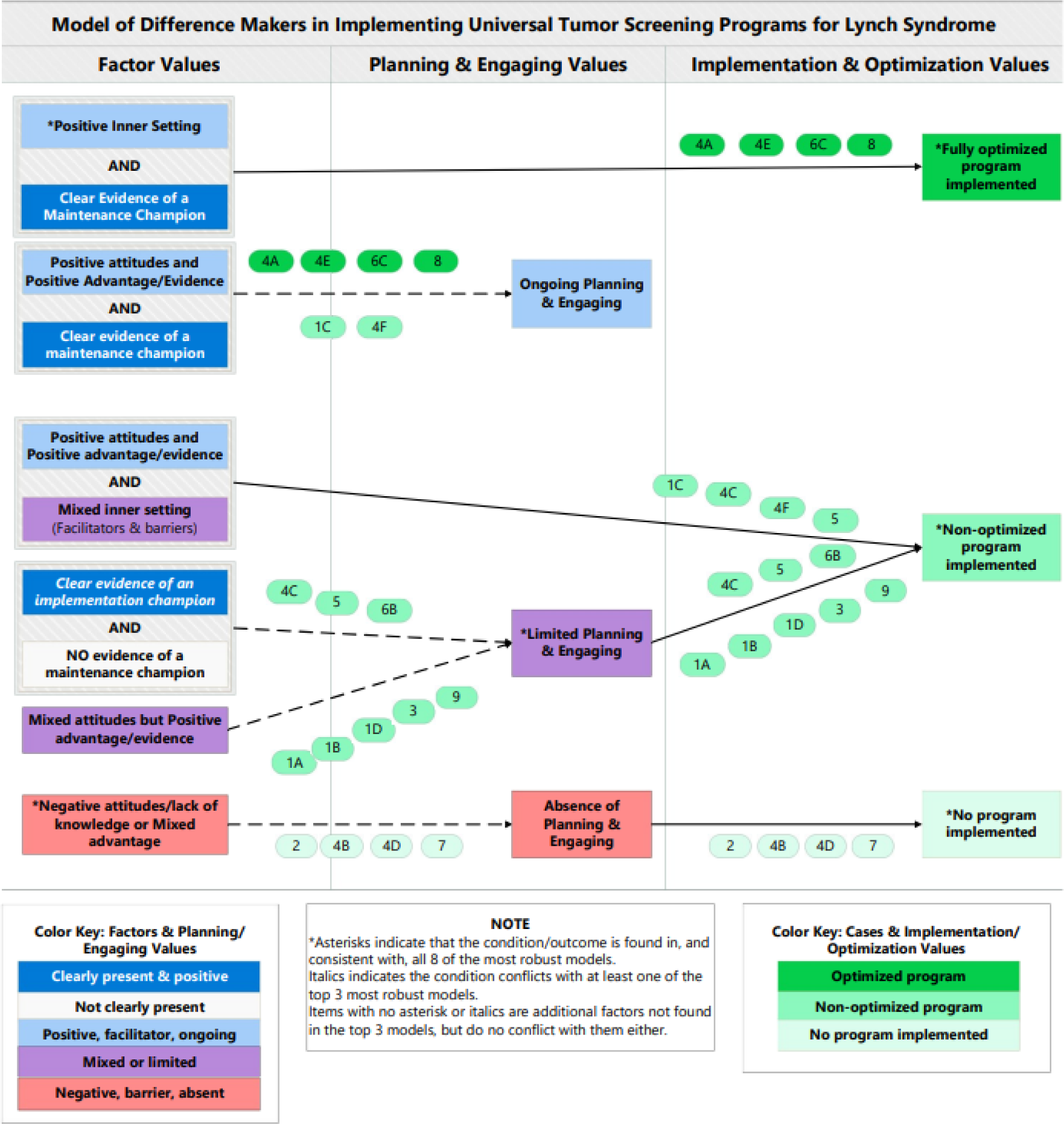
Illustration of the selected CNA model showing which cases fit the patterns of conditions that make a difference for all three ‘Planning & Engaging’ values and all three ‘Implementation & Optimization’ values.

### Factor values differentiating each final outcome value (i.e., optimized, non-optimized, and no UTS program)

#### Optimized UTS Program

According to the model, a distinct combination of two factor values (i.e., conditions) is minimally necessary for fully optimized UTS programs: presence of a maintenance champion and clearly positive inner setting (e.g., sufficient resources, supportive leadership, and solid networks and communication between pathology, oncology, and genetic providers). This combination of conditions distinguishes all four cases with optimized programs (#8, #4E, #4A, #6C) from all other cases.

One role described for maintenance champions in optimized programs is to take charge of quality assurance initiatives as indicated by a genetic counselor maintenance champion at UTS program #6C, *“I am a little bit more involved than my colleagues because I also run the QA so every couple months we ask pathology just to send us a list of all the patients where IHC has been completed and we just double check that we’ve received everything so no one falls through.”* Of note, the two non-optimized programs that also have a maintenance champion present (#1C and #4F) lack a clearly positive inner setting, which is the second condition needed for optimization.

Multiple components can contribute to a positive inner setting at a given organization. For example, tumor boards were described as a useful formal communication network by a genetic counselor at program #8, *“The genetic counselors also go to tumor boards, so [oncologists and pathologists] know we can alert them from the spreadsheet if it looks like somebody has gone through and may not have had a referral.”* Supportive leadership is another valuable inner setting component, as noted by an advanced practice nurse at program #4E, *“[Administrative leaders] were always supportive and onboard, and I think it helped too that our [national leadership] was also encouraging sites to look into [UTS] further…”* Additionally, when asked about the three most important things for successful implementation of a UTS program, a genetic counselor from program #4A responded, *“a physician champion, a passionate genetic counseling staff, and time.”* This illustrates how the availability of sufficient resources [including time allotted to do the job] contributes to a positive inner setting; the quote also reinforces the key role of a champion.

The importance of having both a positive inner setting and a maintenance champion as necessary to be fully optimized is further illustrated by the only other program (#6B) where multiple stakeholders described positive components (and no negative components) of their inner setting, but the program was non-optimized. An oncologist from this program identified the lack of a maintenance champion as the key challenge preventing them from being optimized, *“We don’t have someone who wants to take charge of that either at the genetic side, oncology side, gastroenterology, or pathology side, so we don’t do a post-hoc check of all the patients to make sure we’re not missing anyone.”* Thus, although this program has a positive inner setting, the other condition needed to become fully optimized is clearly missing.

Programs #4F and #1C were similar to the four optimized programs where stakeholders reported *ongoing* planning & engaging. A nurse practitioner at #4F spoke about their implementation process, saying, *“We [implemented UTS] for Commission on Cancer Quality Improvement project; I discussed it with the oncologist and the pathologist and … went back retrospectively and looked at … how the process was working, and we decided as a group to go forward with all colorectal [patients] to be screened by MMR…”* However, this ongoing engagement by itself was insufficient for optimization as evidenced by the lack of other key optimization components at #4F, including a lack of routine reflex testing and referral consistency, and lack of tracking the genetics referrals. In other words, the preferred model provided no evidence that ongoing planning & engaging is minimally sufficient for optimization.

#### Non-optimized UTS program

All 10 non-optimized programs fit at least one of two independent paths of minimally sufficient conditions unique to non-optimized UTS program. Three of these non-optimized programs were mentioned previously because they had only one of the two conditions necessary for optimization. Specifically, program #6B had a positive inner setting, but no maintenance champion; and programs #4F and #1C each had a maintenance champion, but mixed inner setting.

The first path to non-optimization includes the co-occurrence of two difference-making conditions: 1) having knowledgeable stakeholders who hold positive attitudes about UTS and recognize favorable evidence and relative advantages of UTS and 2) a mixed inner setting (e.g. both positive and negative inner setting factors present). The first condition differentiates four of the non-optimized cases (#5, #1C, #4F, #4C) from all cases with no program. Despite the presence of positive attitudes and recognition of advantages of UTS, the four programs that fit this path to non-optimization exhibited a mixed inner setting, which differentiates them from all optimized programs. The mixed inner setting of program #4F is evidenced by stakeholder descriptions of both negative inner setting components (including UTS not remaining among the pathologists’ highest priorities), and positive inner setting components (including UTS remaining a high priority for a nurse practitioner champion) who stated: “*When I get a lull in reports I always call down [to pathology] and say ‘are we still doing this?’ and usually it’s ‘yes’…they just haven’t gotten the report to me*.”

The second path suggests that *limited* planning & engaging may be an intermediate outcome because it is the single difference-maker on the second path to a non-optimized program. A total of eight cases had evidence of limited planning & engaging, including two of the cases that were also consistent with the first path. Planning & engaging was limited for most non-optimized programs simply because it was not ongoing, as illustrated by a genetic counselor at program #5, *“Actually, when we were implementing it, yes, there were meetings across different departments, but once it was initiated, we have not, at least not that I’m aware of.”* Conversely, for program #4C, limited planning & engaging seemed to be due to limited access to a particular key stakeholder type, as reported by a pathologist, *“Unfortunately, we don’t have great access to genetic counselors. [Administration] just simply haven’t put forth the consistent support to have them in place so that we have a direct referral route for a genetic counselor for that visit.”* This quote demonstrates how limited engagement with genetic counselors prevented the consistency of referrals, which is necessary to be fully optimized.

#### No Program

All four cases without a UTS program had no evidence of UTS planning & engaging, which is critical for implementation. This was noted in a quote from a medical director at an organization with no program (#7), *“…making a decision that we are going to do [UTS] really rests with the people with the most expertise. With the oncologists, the surgical oncologists saying this is something that we need to do and we need to implement, and then once that has been decided, it would end up going through our practice guidelines committee, and we would all endorse that.”* This quote reveals a clear mechanism to engage stakeholders at that organization, but the key stakeholders had not decided UTS was necessary and therefore no planning occurred. A physician leader at another health system (#2) noted how challenging it would be to successfully engage stakeholders or agree on a plan, *“How would you get agreement among these providers that are not employees [of the health care system]? Like I say, it’s like herding cats. So, in an integrated environment that’s one challenge of trying to get agreement even among all the different stakeholders. Now when you take it outside an integrated environment, it’s an order of magnitude more difficult.”*

### Conditions differentiating three levels of planning & engaging (i.e., ongoing, limited, and no planning & engaging)

#### Ongoing planning & engaging

The selected CNA model shows a combination of two conditions that constituted the only path leading to ongoing planning & engaging. These conditions include clear evidence of a maintenance champion along with knowledgeable stakeholders who hold positive attitudes about UTS and recognize evidence/advantages favoring UTS, as illustrated by a clinical oncology services director at program #4A:

> *“I think it’s been a strong GI pathology lead, leading it and being the champion of it. It’s made a huge difference. We had several other physicians that understood it and wanted to proceed with it, so I think that was very helpful…but I really think the success is based on the pathologist’s willingness to lead and bring us to where we are at this point.”*

#### Limited Planning & engaging

The selected model provided evidence of a complex ‘causal chain’ whereby two paths lead to limited planning & engaging, and this intermediate outcome subsequently results in a non-optimized program. The first of the two paths to limited planning and engaging consists of the presence of an implementation champion in conjunction with the absence of a maintenance champion, as identified in programs #4C, #5, and #6B. Although an implementation champion can serve as the maintenance champion, this was not always the case (even at optimized sites), and all UTS programs lacking a maintenance champion had limited planning & engaging, as noted by an oncologist at UTS program #6B, *“We don’t track [patients] in any systematic way. We’ve tried off and on [to engage people in tracking] but nobody has taken the lead on that.”*

The second (alternative) path to limited planning & engaging was characterized by a lack of knowledge about UTS among some health system stakeholders, despite all health system stakeholders holding positive perceptions of evidence and relative advantage.

#### No Planning & engaging

The selected model contains another ‘causal chain’ whereby negative views about the evidence for and advantages of UTS among some stakeholders or a lack of stakeholder knowledge about UTS leads to a lack of planning & engaging, which ultimately prevents UTS implementation. The importance of ensuring system stakeholders recognize the evidence for and relative advantage of UTS to initiate engagement was noted by a medical director at health system #7, *“If [GI and Oncology] said, listen, this is something where here is best practice and here is cost-effectiveness and this is something which is universally recommended and we are not doing it, we need to get around to figure out how to implement it… they would bring that through clinical practice guidelines committee and we would, once adopted and approved, get the systems in place to implement.”* Not all stakeholders at sites without a program failed to see the relative advantages of UTS. For example, one physician leader from health system #2A acknowledged the relative advantages of UTS, but lacked an understanding of UTS process, sensitivity, and specificity, which appeared to contribute to concerns and negative attitudes:

> *“We realize that universal [tumor] screening may pick up people that we haven’t previously identified that are at risk for Lynch syndrome, but on the other side, it also may create a lot of false positives that would increase utilization of services. Meaning would we be doing more frequent colonoscopy screenings than are really required and what impact would that have on access to care? So, that’s one of the concerns.”*

This health system stakeholder failed to understand that, although a subset of individuals without Lynch syndrome will have dMMR, subsequent reflex testing using *BRAF* or hypermethylation rules out most of these individuals who do not need genetic counseling and germline testing. Reflex testing is a necessary component of an optimized program because it increases the efficient use of genetic counseling and testing resources to filter out individuals whose cancer is likely of sporadic origin, thereby reducing use of these resources as well as other downstream services, such as increased colonoscopies, by these individuals. This same provider went on to acknowledge that they are also unaware of which stakeholders to engage in discussions about UTS, “…*you’d have to have representatives involved…you know, I’m not sure who would be the best people to be discussing that.”*

### Other models

Although the selected model in Figure 3 was overwhelmingly favored by the research team, there were multiple CNA models that fit the underlying data at varying consistency and coverage thresholds (see supplemental materials which compare the top 9 most robust models). Two key points are worth noting about these other models. First, all models share commonalities and include most of the same factor and outcome values as the selected model. However, most of the other models failed to explain all three values for planning & engaging and/or were missing additional conditions that may be important difference makers based on experience and literature. Second, some models do not support planning & engaging as an intermediate outcome. For example, unlike our selected model, other models suggest that negative views about the evidence for and advantages of UTS among some stakeholders or a lack of stakeholder knowledge about UTS is a “common cause” meaning that it results in both a lack of planning & engaging as well as no UTS program. As another example, one of the other models suggests that ongoing planning & engaging together with a positive inner setting leads to optimized programs as part of a causal chain that was not part of the selected model.

### Other factors not identified as difference makers

A few other factors identified as barriers or facilitators of program implementation by some stakeholders in our study or at certain organizations were not key difference makers in any of the robust CNA models. Cost was a central issue that came out in every interview for health care system #2 (no UTS program) and appeared to influence the overall negative attitude of some stakeholders in the system. However, for all other health care systems and UTS programs in our sample, cost was either discussed in terms of the known cost-effectiveness of UTS or as a prior concern that was overcome by decision-makers. Likewise, although CFIR constructs of peer pressure and cosmopolitanism were found in the qualitative data as a motivation for UTS implementation or ongoing quality assurance, it was not a key difference maker in any of the robust models.

## DISCUSSION

The IMPULSS study was designed to evaluate UTS program implementation and optimization as a use case for understanding system-level implementation of complex programs over multiple service lines to better guide implementation, optimization, and improved patient outcomes.^25^ This study addresses a significant evidence gap by utilizing CNA to systematically compare health care systems at various stages of UTS program implementation to identify key factors that consistently contribute to successful implementation and optimization. The cross-case design assessing multi-level factors revealed clinician-level factors that appear to play a key role in whether health care system stakeholders engage and plan to implement a UTS program, while organization-level factors (e.g., inner setting) were identified as critical for optimization of UTS programs.

Evidence strongly supports the importance of certain components of our results as difference-makers given these conditions and outcome values are present in all the most robust CNA models. Specifically, all models found a positive inner setting (with no negative inner setting components) consistently distinguished fully optimized programs from nearly all non-optimized programs. Additionally, limited planning & engaging made a difference for most non-optimized programs in all robust models. Finally, negative attitudes and lack of knowledge or mixed perceptions of the relative advantages of UTS over other Lynch syndrome identification approaches at the clinician-level was a key difference maker for the absence of planning & engaging and/or absence of a program in all models. However, results are less clear as to whether this condition acts directly to prevent UTS program implementation or acts indirectly (as the first part of a causal chain) whereby it prevents stakeholders from planning & engaging, which in turn contributes to no program (as the second part of a causal chain).

Results of this study were used to develop an interactive, web-based toolkit consisting of information, decision tools, planning guides, and resources targeted to 4 phases of UTS program implementation: 1) Deciding to implement, 2) planning how to implement, 3) improving (i.e., optimizing) an existing program, and 4) considering different methods to identify LS for those with existing programs. Based on CNA results, organizational-level decision-making and planning involve multiple decisions by various health care system stakeholders during different phases of implementation. In the deciding phase, (for those with no program) system stakeholders must be educated to increase knowledge and accept the evidence and relative advantages that favor LS screening of all colorectal and endometrial tumors. In the implementation planning phase, we want to facilitate successful engagement of all stakeholders and ensure planning for an optimized program. Those with an existing, but non-optimized program, may need to continue ongoing planning, improve aspects of their inner setting (such as communication networks), engage a maintenance champion, and/or ensure all key stakeholders hold positive attitudes toward UTS.

Our results align well with other studies that identified factors important for successful implementation of genomic medicine in a single system or in multiple health care systems. Qualitative analysis of system-level barriers from IGNITE study teams found negative attitudes, lack of knowledge, and lack of buy-in contributed to lack of implementation across research programs^31^, and in 2021 the network identified four implementation strategies in common across six genomic medicine studies. The four strategies identified (i.e., obtaining and using stakeholder feedback, identifying early adopters, conducting education meetings, meeting with an expert)^32^ are consistent with the lack of planning and engaging or limited ongoing planning and engaging that differentiated health care systems with no UTS or non-optimized programs from optimized UTS programs in our model. However, our select model also identified additional conditions needed for successful and ongoing engagement (e.g., a maintenance champion).

### Strengths and Limitations

This study illustrates the value of multi-value CNA, supported by robust qualitative data, as a novel approach to uncover complexity that arises when more than one independent path leads to the same intermediate outcome and to reveal support for causal chains, whereby intermediate outcome values lead to final outcomes. Nevertheless, given model ambiguity it is possible that the same conditions may be contributing to both the intermediate and final outcomes as part of a common cause, rather than causal chain. Typically, whenever a causal chain is identified in CNA, there is also a corresponding common cause model (although the reverse is not true - there is not always a corresponding causal chain model found).

Although the selected model may have been overfit, our exhaustive qualitative data and theoretic guidance by the CFIR supported the additional “pathway” that explained two additional cases and it was more important to identify all possible difference makers so they could be addressed in our toolkit. Given that the underlying qualitative data were based on available key stakeholders collected at a single point in time, this limits the ability to observe changes in planning and engaging that may occur over time. This may explain why our preferred model did not identify ongoing planning & engaging as an intermediate outcome on the path to program optimization. Longitudinal assessment of UTS programs in real-time would be necessary to clarify and confirm the importance of ongoing planning & engaging to the final outcome of optimization. Additionally, although we combined the inner setting components into a theory construct-level difference maker, every organization is unique and there may be different aspects of the inner setting to be addressed using different strategies in order to become fully optimized at any given organization.

This study was successful at uncovering multi-level difference makers for implementation of a complex intervention (LS UTS program) within multiple health care systems. However, CFIR factors that make a difference for simple / non-complex interventions might vary and there may be different facilitators and barriers when implementation is focused on adoption at a clinician level (rather than the system level as is required for UTS). Finally, this study is based on CFIR 1.0. as CFIR 2.0 was not available until cross-case coding and matrix heat mapping were completed. However, as we reviewed qualitative data to support the model, we believe it remains relatively consistent with CFIR 2.0.

### Conclusions and Future Directions

The IMPULSS study provides a model to understand program implementation in health care systems using LS UTS program implementation as a use case. We have created an interactive toolkit for LS UTS implementation and optimization based on the data from this study that resides on the Lynch Syndrome Screening Network (LSSN) website (www.lynchscreening.net).^33^ Importantly this toolkit includes not only primary implementation guidance but tools for ongoing assessment and optimization. In our study, non-optimized programs were missing two or more of the 5 optimization factors at the time of data collection; however, several had the potential to become optimized, and focusing on difference makers for optimization (rather than those required for initial implementation) using the toolkit may provide helpful guidance. While the interactive toolkit on the LSSN website contains information and decision tools specific to LS UTS programs, implementation strategies from the ERIC compilation^34^ could be mapped to our selected CNA solution to create a general guide for organizational implementation of any similarly complex program requiring coordination across multiple service lines and for innovations with multiple program design options. Finally, findings from this study, together with the connected strategies could be tested to determine prospectively whether they help with implementation and optimization of UTS or other precision health programs.

## Data availability

available on request

## Acknowledgements

We gratefully acknowledge all our project managers and student trainees without whom a project of this scope and complexity would not have been possible. The collaborations of health care systems participating in this study are possible through the Health care System Research Network (HCSRN) and the Cancer Research Network (CRN). Additional guidance on study activities and dissemination is provided in collaboration with the Lynch Syndrome Screening Network (LSSN). Finally, the authors thank our health system stakeholders who made this work possible.

## Author Contributions

Conceptualization: all authors; Funding Acquisition: AKR; Data Acquisition: all authors; Data analysis: DC, ZS, JS, AKR; Writing-original draft: DC, ZS, AKR; Writing-review & editing: all authors

## Ethics Declaration

This study was approved by the Geisinger IRB (#2017-0238), which served as the central IRB (cIRB) for all sites except Sutter Health, and the Sutter Health IRB (#2017.134EXP).

## CONFLICT OF INTERST STATEMENTS

none declared

