## supplemental for "Using Coincidence Analysis to Identify Causal Chains of Factors Associated with Implementation and Optimization of Lynch Syndrome Tumor Screening Across Multiple Health care Systems"

**Supplemental Methods Appendix**

### Methods

***CNA overview***

Coincidence analysis (CNA) is a type of configurational comparative method (CCM) that uses R software packages to conduct cross-case comparisons of factor and outcome values. CNA is based on a bottom-up algorithm that identifies patterns of factor values (a.k.a., conditions) that are minimally sufficient for an outcome value to occur (Baumgartner & Ambuhl, 2023). Unlike most traditional analytic approaches, CCMs (such as CNA) do not rely on correlations between variables. Instead, CNA reveals patterns that occur regularly in calibrated data. Given that the underlying goal of CCMs is to gain a deeper understanding of the data, the analytic approach is iterative. As with qualitative analysis, data used for CNA are often re-coded as additional insights come to light; and it is common to refine the way in which data are calibrated. Below we describe details about how we calibrated and consolidated data, conducted CNA, and selected from among multiple models that were output by the software.

***Data consolidation and re-calibration process***

A data matrix of select factors had been previously consolidated and selected for use in CNA as described elsewhere (Salvati et al., 2023) and summarized in Supplemental Figure 1.

**Supplemental Figure 1**. Data matrix heat map showing CFIR factors hypothesized to make a difference for the main outcome.

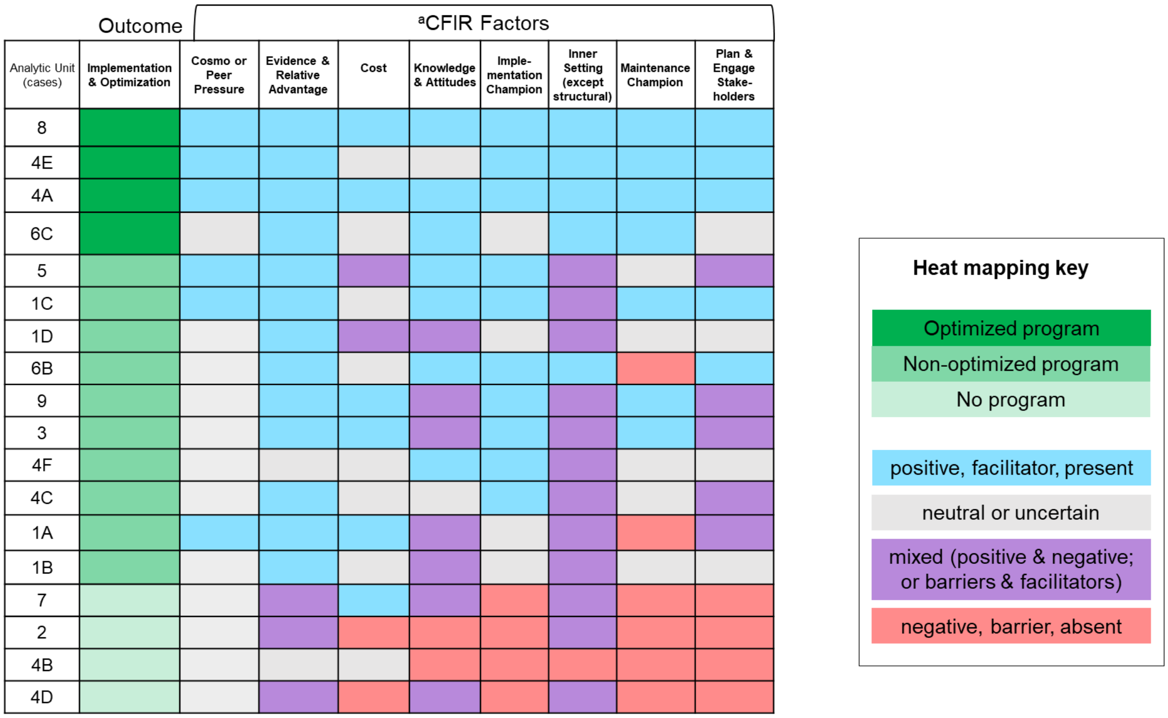


However, further consolidation of factors and factor values was necessary to reduce data fragmentation. Data fragmentation is problematic in CNA because it can lead to false positive findings. Thus, to help reduce data fragmentation, further consolidation and re-calibration were completed through the following steps:

- Dichotomizing three factors (implementation champion, maintenance champion, and cosmopolitanism or peer pressure) as clearly present and positive = 1 versus not clearly present and positive= 0.
- Combining external networks together with peer pressure because they both are influences from outside the organization that may have a positive impact on UTS implementation. Specifically, cases with either of these factors present were assigned a value of 1 and all others were assigned a value of 0 for this newly combined factor.

- Combining perceptions of UTS evidence and relative advantage with attitudes and knowledge such that all positive and no mixed or negative=4, positive evidence/advantage but mixed attitudes/knowledge=2, and mixed evidence/advantage or negative attitudes/lack of knowledge =1.
- Assigning an inner setting value of 2 to the only case with a negative inner setting
- Dichotomizing cost as the presence of at least some cost concerns = 1 versus no cost concerns= 0.
- Redefining 'planning and engaging' to equate to the presence of ongoing planning and engaging among all key stakeholders as opposed to any evidence of stakeholder planning and engaging.

Notably, redefining and then re-calibrating 'planning and engaging' was done, not only to reduce fragmentation, but also because the team believed that the ongoing nature and engagement of all stakeholders was more likely to contribute to UTS program optimization rather than planning/engaging that was either limited to when the program was first initiated or included only some key stakeholders (e.g., genetics was no longer engaged in ongoing planning). To ensure accuracy when re-defining this factor, all text from interview transcripts previously coded as either planning or stakeholder engagement was reviewed for all cases and the entirety of the original stakeholder transcripts were reviewed in cases that had previously been color-coded as non-salient/neutral. This resulted in three total values for the 'planning and engaging' factor (i.e., ongoing planning and engaging =4, limited planning and engaging = 2, and no planning and engaging = 1).  The final consolidated and calibrated data matrix is shown in Supplemental Figure 2.

**Supplemental Figure 2.** Final consolidated and calibrated data matrix of factors and implementation outcomes.

| | CFIR Factors combined | | | | | | Implementation Outcomes | |
|---|---|---|---|---|---|---|---|---|
| Cases | Cosmopolitanism or Peer Pressure | Imple-mentation Champion | Maintenance Champion | Cost Concerns | Evidence, advantage, knowledge, attitudes | Inner Setting (except structural) | Initial and Ongoing Planning & Engaging of Stakeholders | Implementation & Optimization |
| 8 | 4 | 4 | 4 | 0 | 4 | 4 | 4 | 2 |
| 4E | 4 | 4 | 4 | 0 | 4 | 4 | 4 | 2 |
| 4A | 4 | 4 | 4 | 0 | 4 | 4 | 4 | 2 |
| 6C | 0 | 0 | 4 | 0 | 4 | 4 | 4 | 2 |
| 5 | 4 | 4 | 0 | 1 | 4 | 2 | 2 | 1 |
| 1C | 4 | 4 | 4 | 0 | 4 | 2 | 4 | 1 |
| 1D | 0 | 0 | 0 | 1 | 2 | 2 | 2 | 1 |
| 6B | 0 | 4 | 0 | 0 | 4 | 4 | 2 | 1 |
| 9 | 0 | 4 | 4 | 0 | 2 | 2 | 2 | 1 |
| 3 | 0 | 4 | 4 | 0 | 2 | 2 | 2 | 1 |
| 4F | 0 | 4 | 4 | 0 | 4 | 2 | 4 | 1 |
| 4C | 0 | 4 | 0 | 0 | 4 | 2 | 2 | 1 |
| 1A | 4 | 0 | 0 | 0 | 2 | 2 | 2 | 1 |
| 1B | 0 | 0 | 0 | 0 | 2 | 2 | 2 | 1 |
| 7 | 0 | 0 | 0 | 0 | 1 | 2 | 1 | 0 |
| 2 | 0 | 0 | 0 | 1 | 1 | 2 | 1 | 0 |
| 4B | 0 | 0 | 0 | 0 | 1 | 2[a] | 1 | 0 |
| 4D | 0 | 0 | 0 | 1 | 1 | 2 | 1 | 0 |

[a]Inner Setting was calibrated negatively for this case but was combined with "mixed/limited" cases to reduce data fragmentation.

**Data Calibration of Factors/Outcomes**

- 4=Clearly present & positive
- 1=Clearly present & negative
- 0=Not clearly present

- 4=positive, facilitator, ongoing
- 2=mixed or limited
- 1=negative, barrier, absent

- 2=Optimized program
- 1=Non-optimized program
- 0=No program

Each row depicts a deidentified case (first column), showing the combination of CFIR factor values (i.e., conditional configuration) and respective implementation outcome values associated with that case.

#### *Conducting CNA and robustness testing*

The calibrated data matrix along with the CNA and frscore packages in R statistical software were used to conduct multi-value coincidence analysis (mv-CNA) (Baumgartner and Ambuhl, 2023). Factors were ordered based on theory (Figure 1) so that downstream outcomes are not allowed to cause the preceding (upstream) factors or hypothesized intermediate outcome values. However, ordering does not stipulate any relationships (Baumgartner and Epple, 2013), meaning that such relationships only show up in CNA models if they are substantiated by the underlying data. In the ordering statement the parameter "strict" was set to "FALSE" which meant that the CFIR constructs in Supplemental Figure 3 would not be modeled as causes of each other.

**Supplemental Figure 3**. Hypothesized factors that may make a difference for implementation outcomes.

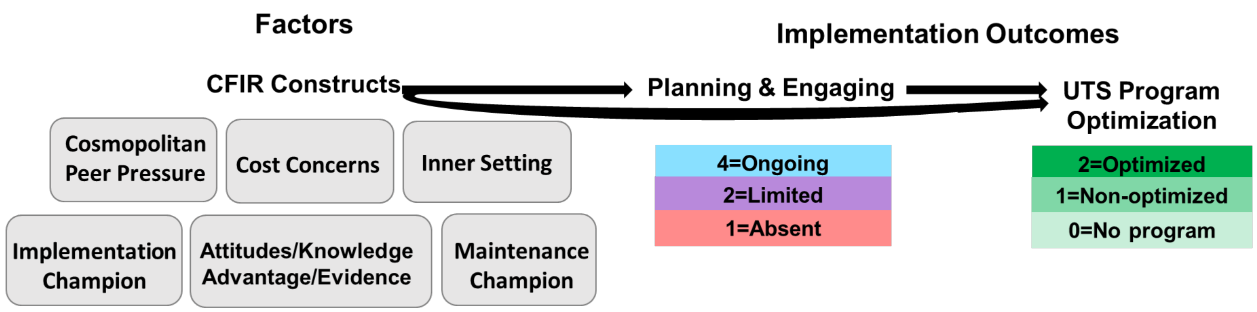


Consistency and coverage thresholds for CNA were initially set to .90. Consistency and coverage are used by the CNA software to build models and serve as key measures to consider when multiple models are output by the software. Consistency is analogous to positive predictive value and coverage is analogous to sensitivity. Although consistency and coverage scores of 1.0 indicate perfect model fit, it is possible for such models to be overfit, meaning that they may include additional factor values that are not truly difference makers. Consequently, it can be useful to review models that are output at different consistency and coverage thresholds and identify commonalities across models based on submodel/supermodel relationships used to assess relative fit robustness (Parkkinen & Baumgartner, 2020). The frscore package in R was used to run models at various consistency and coverage thresholds between .75 and 1.0 and identify models with the highest relative fit robustness scores. Other parameters were set as follows: 'scoretype' ='full', 'score normalization' = truemax, and 'maxsols' = 600. The CNA code used is shown below.

```
>
>
> library(frscore)
>
> frscored_cna(subsetFinal, type = "mv",
+              n.init = 20,
+              comp.method="is.submodel",
+              fit.range = c(1, 0.75), granularity = 0.05,
+              normalize = "truemax", maxsols=600,
+              ordering = "PlanEngage < Optimize",
+              strict = TRUE,  details=TRUE)
```

**Model Output**

There was substantial model ambiguity in both the initial modeling and fit robustness modeling as is often the case when there are multiple conditions and outcomes along with multiple values for each. Model ambiguity means that there is more than one model that fits the underlying data at various consistency and coverage thresholds.

When there is model ambiguity the most robust models are often reviewed for commonalities. For example, all of these models explained all three values of the final outcome (OPTIMIZE). Additionally, all models include positive inner setting (INNERSET=4) as a difference-making condition for fully optimized programs (i.e., OPTIMIZE=2). There were, however, some differences across models; and only models #4 and #7 explained all three values for the hypothesized intermediate outcome of planning and engaging (PLANENGAGE).

```
FR-scored reanalysis series with fit range 1 to 0.75 with granularity 0.05
Score type: full || score normalization: truemax
maxsols set to 600 -- 0 model types excluded from scoring


-----

Model types:

                                                                  outcome
1  OPTIMIZE=0,OPTIMIZE=1,OPTIMIZE=2,PLANENGAGE=1,PLANENGAGE=4
2  OPTIMIZE=0,OPTIMIZE=1,OPTIMIZE=2,PLANENGAGE=4
3  OPTIMIZE=0,OPTIMIZE=1,OPTIMIZE=2,PLANENGAGE=1,PLANENGAGE=2
4  OPTIMIZE=0,OPTIMIZE=1,OPTIMIZE=2,PLANENGAGE=1,PLANENGAGE=2,PLANENGAGE=4
5  OPTIMIZE=0,OPTIMIZE=1,OPTIMIZE=2,PLANENGAGE=1,PLANENGAGE=2,PLANENGAGE=4
6  OPTIMIZE=0,OPTIMIZE=1,OPTIMIZE=2,PLANENGAGE=2,PLANENGAGE=4
7  OPTIMIZE=0,OPTIMIZE=1,OPTIMIZE=2,PLANENGAGE=1,PLANENGAGE=4
8  OPTIMIZE=0,OPTIMIZE=1,OPTIMIZE=2,PLANENGAGE=1,PLANENGAGE=4
9  OPTIMIZE=0,OPTIMIZE=1,OPTIMIZE=2,PLANENGAGE=1,PLANENGAGE=2
```

Note each Boolean solution is followed by a graphical representation of that solution. In the graphs the dots symbolize a conjunction (meaning that more than one condition is necessary for an outcome value).

1 (EVATT=1<->OPTIMIZE=0)*(PLANENGAGE=2<->OPTIMIZE=1)*(INNERSET=4<->OPTIMIZE=2)*(EVATT=1<->PLANENGAGE=1)*(EVATT=4*MAINTCHAMP=4<->PLANENGAGE=4)

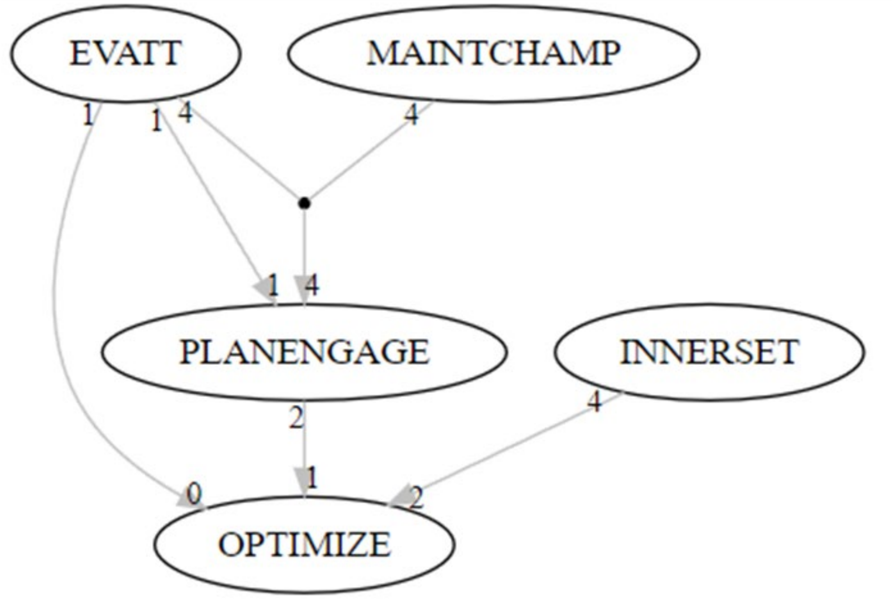


2 (EVATT=1<->OPTIMIZE=0)*(PLANENGAGE=2<->OPTIMIZE=1)*(INNERSET=4*PLANENGAGE=4<->OPTIMIZE=2)*(EVATT=4*MAINTCHAMP=4<->PLANENGAGE=4)

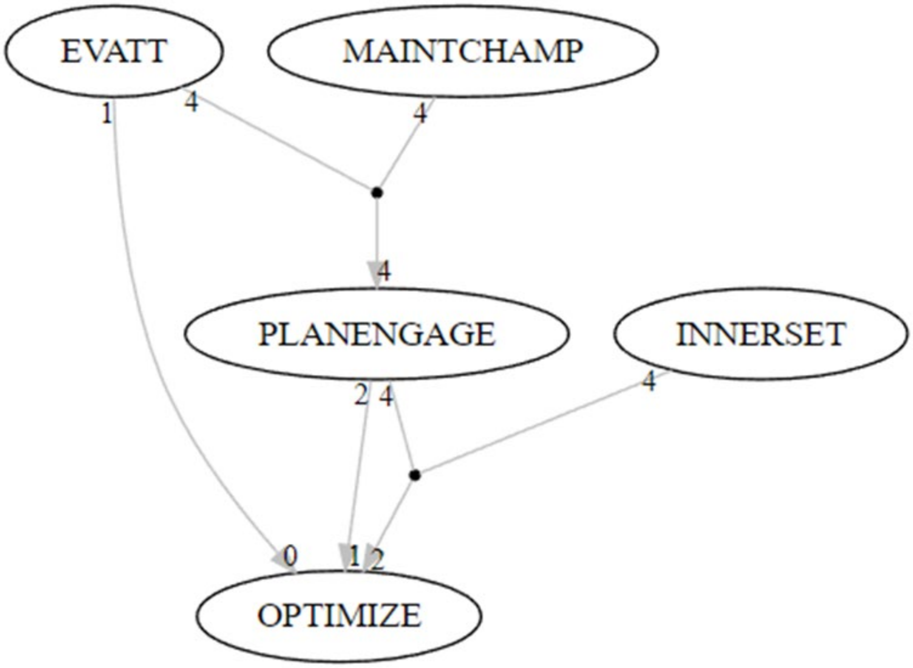

3 (EVATT=1<->OPTIMIZE=0)*(PLANENGAGE=2<->OPTIMIZE=1)*(INNERSET=4<->OPTIMIZE=2)*(EVATT=1<->PLANENGAGE=1)*(EVATT=2+EVATT=4*MAINTCHAMP=0<->PLANENGAGE=2)

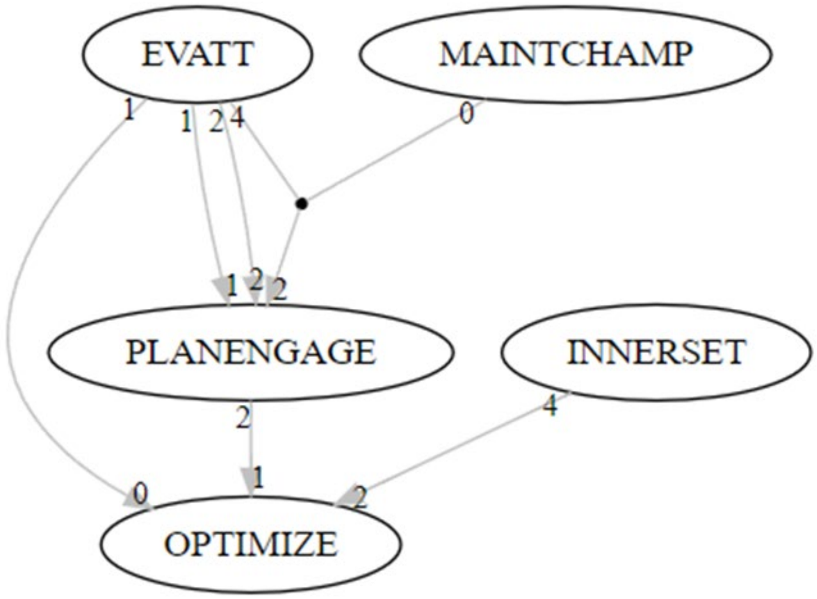


4 (EVATT=1<->OPTIMIZE=0)*(PLANENGAGE=2+EVATT=4*INNERSET=2<->OPTIMIZE=1)*(INNERSET=4*MAINTCHAMP=4<->OPTIMIZE=2)*(EVATT=1<->PLANENGAGE=1)*(EVATT=2+IMPCHAMP=4*MAINTCHAMP=0<->PLANENGAGE=2)*(EVATT=4*MAINTCHAMP=4<->PLANENGAGE=4)

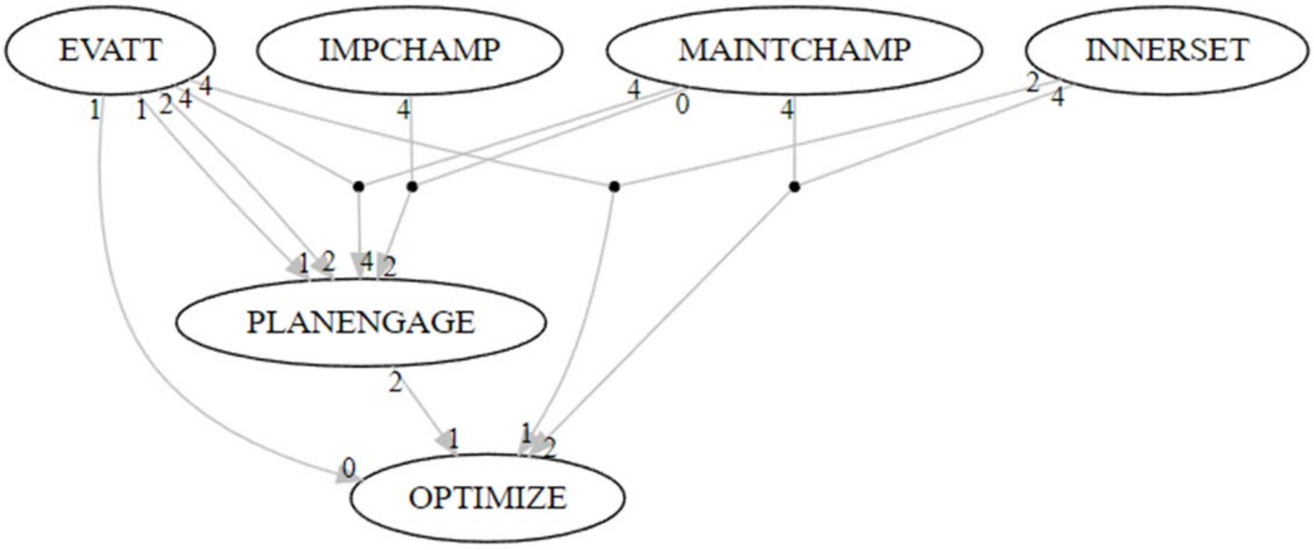

5 (PLANENGAGE=1<->OPTIMIZE=0)*(PLANENGAGE=2+EVATT=4*INNERSET=2<->OPTIMIZE=1)*(INNERSET=4*MAINTCHAMP=4<->OPTIMIZE=2)*(EVATT=1<->PLANENGAGE=1)*(EVATT=2+IMPCHAMP=4*MAINTCHAMP=0<->PLANENGAGE=2)*(EVATT=4*MAINTCHAMP=4<->PLANENGAGE=4)

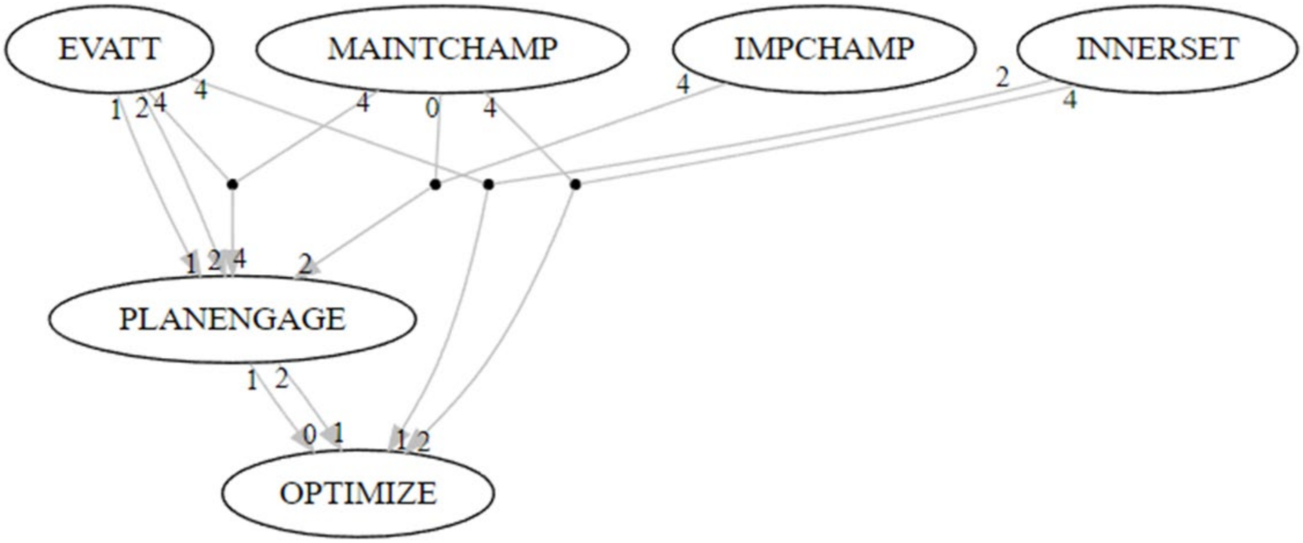


6 (EVATT=1<->OPTIMIZE=0)*(PLANENGAGE=2<->OPTIMIZE=1)*(INNERSET=4<->OPTIMIZE=2)*(EVATT=2+EVATT=4*MAINTCHAMP=0<->PLANENGAGE=2)*(EVATT=4*MAINTCHAMP=4<->PLANENGAGE=4)

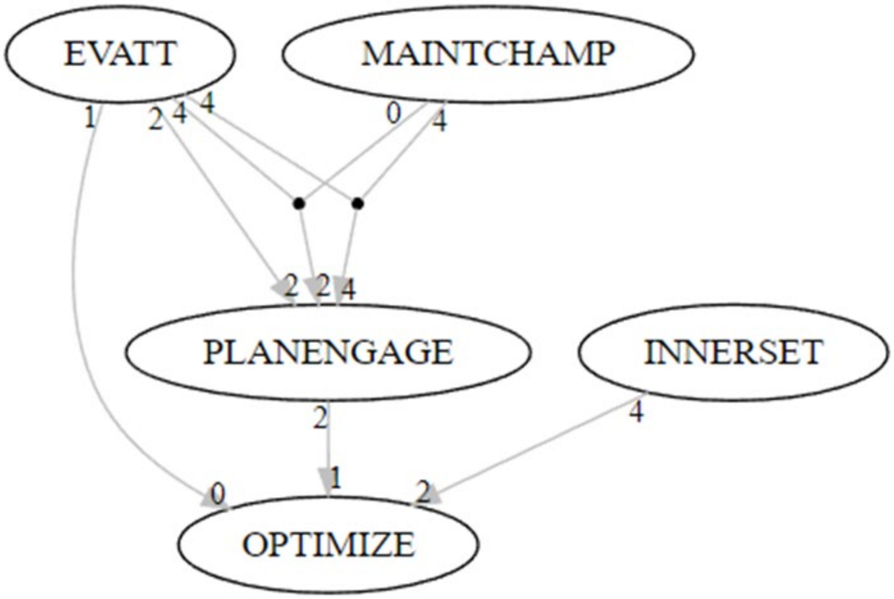

7
(PLANENGAGE=1<->OPTIMIZE=0)*(PLANENGAGE=2<->OPTIMIZE=1)*(INNERSET=4<->OPTIMIZE=2)*(EVATT=1<->PLANENGAGE=1)*(EVATT=4*MAINTCHAMP=4<->PLANENGAGE=4)

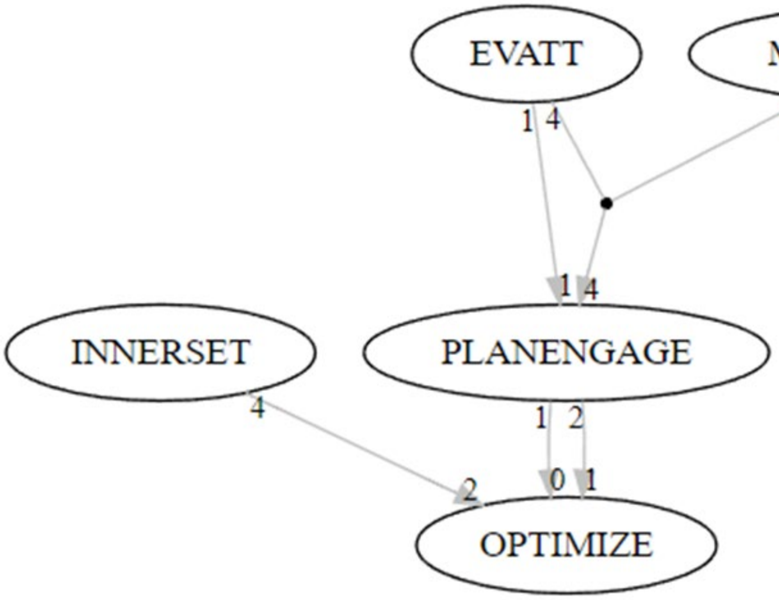


8
(EVATT=1<->OPTIMIZE=0)*(PLANENGAGE=2<->OPTIMIZE=1)*(INNERSET=4*MAINTCHAMP=4<->OPTIMIZE=2)*(EVATT=1<->PLANENGAGE=1)*(EVATT=4*MAINTCHAMP=4<->PLANENGAGE=4)

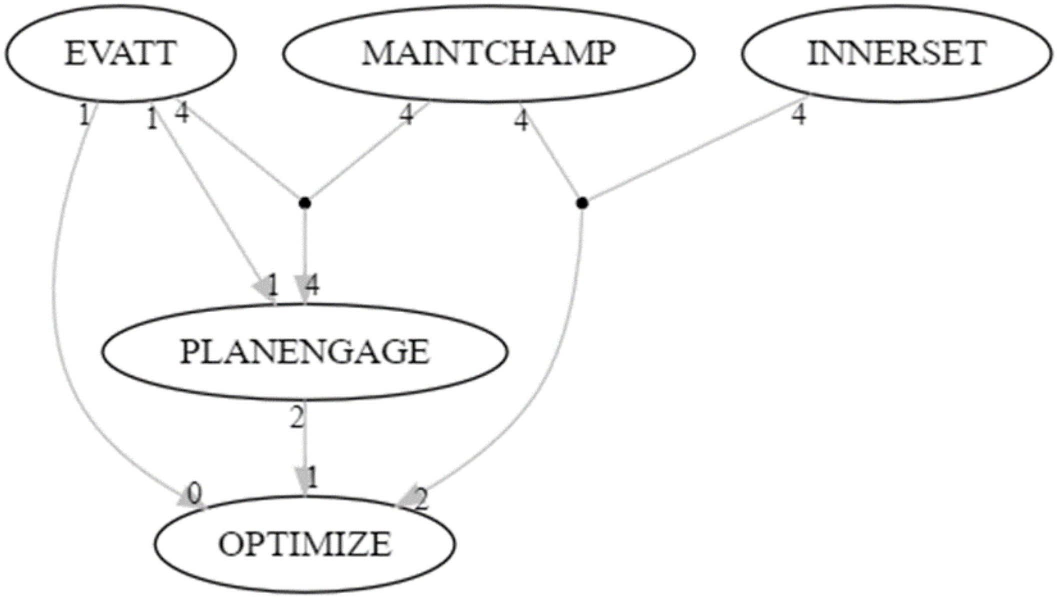

```
9                                                                  (PLANE
NGAGE=1<->OPTIMIZE=0)*(PLANENGAGE=2<->OPTIMIZE=1)*(INNERSET=4<->OPTIMIZE=2)*(EVATT
=1<->PLANENGAGE=1)*(EVATT=2+EVATT=4*MAINTCHAMP=0<->PLANENGAGE=2)
```

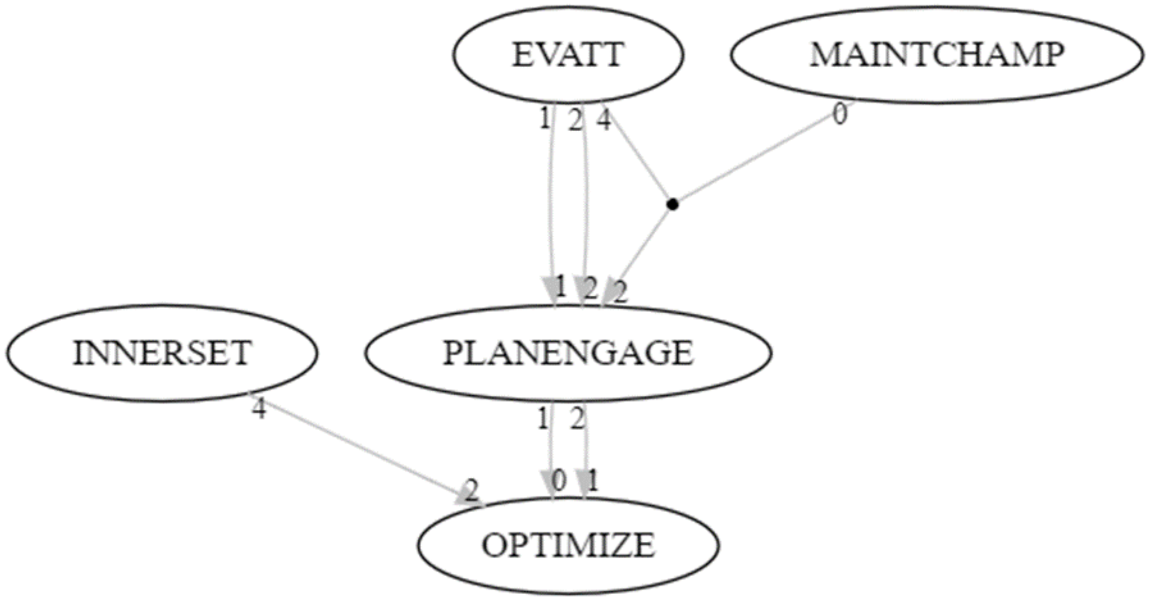


```
   consistency coverage complexity inus exhaustiveness faithfulness coherence
redundant cyclic score tokens norm.score
1         0.80      0.8          6 TRUE      0.8333333    0.7142857 0.8333333
FALSE  FALSE   182       3  1.0000000
2         1.00      0.8          6 TRUE      0.5454545    0.8571429 0.8888889
FALSE  FALSE   170      12  0.9340659
3         0.80      0.8          7 TRUE      0.8333333    0.7142857 0.8333333
FALSE  FALSE   168       3  0.9230769
4         1.00      1.0         12 TRUE      0.5000000    1.0000000 1.0000000
FALSE  FALSE   141      16  0.7747253
5         1.00      1.0         12 TRUE      0.5000000    1.0000000 1.0000000
FALSE  FALSE   126      16  0.6923077
6         0.80      0.8          8 TRUE      0.8333333    0.7142857 0.8333333
FALSE  FALSE   120       3  0.6593407
7         0.80      0.8          6 TRUE      0.8333333    0.7142857 0.8333333
FALSE  FALSE   118       3  0.6483516
8         1.00      0.8          7 TRUE      0.6666667    0.8571429 0.8888889
FALSE  FALSE   105      12  0.5769231
9         0.80      0.8          7 TRUE      0.8333333    0.7142857 0.8333333
FALSE  FALSE   101       3  0.5549451
```

#### Model Selection Process

The top 9 models were illustrated graphically and a key team member reviewed the qualitative data to identify quotes to support the various different models. The models were shared with the larger research team to determine which one made the most sense. Model (#5) was selected unanimously as the preferred model. This preferred model has perfect consistency and coverage and substantiated two causal chains with limited planning and engaging and no planning and engaging as intermediate outcomes along the paths to non-optimization and no program, respectively. Six of the 9 most robust models identified one or more causal chains in which one or more planning & engaging values are an intermediate outcome. However, common cause models were also identified in which the same condition directly and independently impacted a value for both ‘planning & engaging’ and ‘implementation or optimization’ (rather than as part of a causal chain).

The preferred model was more complex and may be overfit, but it was important to identify all key factors that make a difference for implementation and optimization so that these can be addressed in the toolkit and future interventions. Notably, model 4 was almost the same as the preferred model #5, but showed the common cause whereby negative attitudes/lack of knowledge or mixed advantage led to both a lack of planning and engaging as well as no program implementation.

Ultimately there was a lot of overlap across all models, lending credibility to our findings. Nevertheless, there were a few quotes that could support a couple of other models (such as Model #2 which had a causal chain leading to program optimization). However, the quotes primarily supported the selected model (#5), which created the most coherent story and support for planning and engaging as an intermediate outcome.

https://doi.org/10.1177/0049124113502948

**Acknowledgments**

We would like to thank Michael Baumgartner for his support in helping to interpret some of the complex output and troubleshooting the CNA program. Additionally, we thank Christoph Falk for developing and programming the plot function with which we drew the graphs of the solutions included in this appendix to illustrate the similarities more easily across the most robust models.
